# Application of ARIMA, hybrid ARIMA and Artificial Neural Network Models in predicting and forecasting tuberculosis incidences among children in Homa Bay and Turkana Counties, Kenya

**DOI:** 10.1101/2022.07.07.22277378

**Authors:** Siamba Stephen, Otieno Argwings, Koech Julius

## Abstract

**Background:** Tuberculosis (TB) infections among children (below 15 years) is a growing concern, particularly in resource-limited settings. However, the TB burden among children is relatively unknown in Kenya where two-thirds of estimated TB cases are undiagnosed annually. Very few studies have used Autoregressive Integrated Moving Average (ARIMA), hybrid ARIMA, and Artificial Neural Networks (ANNs) models to model infectious diseases globally. We applied ARIMA, hybrid ARIMA, and Artificial Neural Network models to predict and forecast TB incidences among children in Homa bay and Turkana Counties in Kenya.

**Methods:** The ARIMA, ANN, and hybrid models were used to predict and forecast monthly TB cases reported in the Treatment Information from Basic Unit (TIBU) system for Homa bay and Turkana Counties between 2012 and 2021. The data were split into training data, for model development, and testing data, for model validation using an 80:20 split ratio respectively.

**Results:** The hybrid ARIMA model (ARIMA-ANN) produced better predictive and forecast accuracy compared to the ARIMA (0,0,1,1,0,1,12) and NNAR (1,1,2) [12] models. Furthermore, using the Diebold-Mariano (DM) test, the predictive accuracy of NNAR (1,1,2) [12] versus ARIMA-ANN, and ARIMA-ANN versus ARIMA (0,0,1,1,0,1,12) models were significantly different, p<0.001, respectively. The 12-month forecasts showed a TB prevalence of 175 to 198 cases per 100,000 children in Homa bay and Turkana Counties in 2022.

**Conclusion:** The hybrid (ARIMA-ANN) model produces better predictive and forecast accuracy compared to the single ARIMA and ANN models. The findings show evidence that the prevalence of TB among children below 15 years in Homa bay and Turkana Counties is significantly under-reported and is potentially higher than the national average.

## Introduction

### Background

Tuberculosis (TB) is a highly infectious disease ranked among the top ten most lethal causes of mortality. Approximately 33% of the global population has been plague-ridden with TB, particularly in developing countries [1]. In 2016, over 10 million new TB cases were reported globally with children below 15 years of age accounting for about 7% of those cases [3]. In 2018, about 1 million TB disease cases and over 230,000 TB-related deaths occurred among children below 15 years with about 55% of these reported TB cases going undiagnosed and/or unreported [2].

Pediatric TB is usually overlooked [4] amid diagnosis and treatment challenges. Developing countries account for over 85% of new cases of TB globally with Asian and African countries contributing 61% and 25% of global new TB cases [2] respectively. In 2016, approximately 7 countries, globally, accounted for close to 65% of all new TB cases [2] while in 2019, 30 high TB burdened countries accounted for 87% of all new TB cases while only 8 countries accounted for approximately 67% of the total new TB cases [5].

The TB burden in Sub-Saharan Africa (SSA) is far much greater and is exacerbated by poverty, political strive, and weak health systems which have curtailed the implementation of TB control interventions. Consequently, TB has become an enormous burden to health systems that are already overstretched [6].

Tuberculosis is a disease of major concern in Kenya and is among the top five causes of mortality. Kenya is listed among the top 30 TB high burdened countries [7]. Kenya is also among 14 countries globally that suffer from the TB, TB-HIV co-infection, and Multi-Drug Resistant TB [8] triple burden. The TB prevalence for Kenya in 2015 was 233 per 100,000 population with a mortality of 20 per 100,000. In Kenya, the TB case notification increased from 11,000 to 116,723 between 1990 and 2007 [9] occasioned by the HIV epidemic and improved case detection due to improved diagnostic capacity.

The use of mathematical models in the modeling of epidemic interactions within populations has been detailed extensively. While existing interventions to control TB have been partially successful, within the context of resource constraints, mathematical modeling can increase understanding and result in better policies toward the implementation of effective strategies that would compound better health and economic benefits [10]. In addition, mathematical models are essential in guiding policymakers in resource allocation toward the prevention and control of diseases.

Several studies have utilized ARIMA, Seasonal ARIMA (SARIMA), neural network, and hybrid ARIMA models to model TB incidences in other countries [11, 13] and in these studies, the hybrid models were demonstrated to offer better predictive and forecast accuracy. In Africa, Azeez *et al*. compared the predictive capabilities of the SARIMA and the hybrid SARIMA neural network auto-regression (SARIMA-NNAR) models in modeling TB incidences in South Africa and the SARIMA-NNAR model was found to have better goodness-of-fit [12]. In addition, Li *et al*. compared the the predictive power of the ARIMA and ARIMA-generalized regression neural network (GRNN) hybrid models in forecasting TB incidences in China and concluded that the hybrid model was superior to the single ARIMA model [13].

The ARIMA and Neural Networks models have also been applied in modeling other infectious diseases. Zeming and Yanning predicted HIV-AIDS incidences in China in 2017 using the back propagation (BP) artificial neural network and the ARIMA models and concluded that the hybrid (BP-ANN) model offered better predictive power compared to the single ARIMA model [14]. Zhou *et al*. modeled the prevalence of schistosomiasis in Qianjiang city in China using a hybrid model of ARIMA-NARNN (Nonlinear Autoregressive Neural Network) and concluded that the hybrid ARIMA-NARNN model produced high-quality prediction accuracy [15]. Yu *et al*. used the hybrid seasonal ARIMA and NARNN model to forecast incidences of the Coxsackie viral infection in Shenzhen China, and concluded the hybrid seasonal ARIMA-NARNN was the best model [16].

While hybrid ARIMA models have been applied in forecasting both the short-term and long-term incidences of infectious diseases in other countries, there has been little to no application of these cutting-edge methods in African countries with the majority of the models limited to only ARIMA models [17]. In Kenya, while ARIMA models have been applied in forecasting disease incidence [18], very little has been done in the application of hybrid ARIMA models in predicting disease incidence except in non-public health settings such as agriculture and economics.

The popularity of ARIMA models stems from their flexibility to represent varieties of time series with simplicity but with a profound limitation stemming from their linear assumptions which in many cases is usually impractical [19] since real-world applications mainly involve data exhibiting non-linear patterns. Consequently, to overcome this disadvantage, non-linear stochastic models such as the ANN models have been proposed [20]. Despite this, a single ANN model is not able to incorporate both linear and non-linear patterns and this has led to the adoption of hybrid models that can address this challenge [21]. To attain a higher degree of predictive and forecasting accuracy, theoretical and empirical findings show that combining different models can be effective [22].

To better understand the status of TB infection among children in Kenya, it is important to assess the trend and forecast these incidences using available surveillance data and novel models to elicit a better understanding and innovative interventions to curtail the spread of pediatric TB in Kenya. This study compares linear-based ARIMA, non-linear-based ANN, and hybrid ARIMA in modeling TB incidences among children below years in Homa bay and Turkana counties in Kenya.

## Materials and methods

### Study design and setting

This was a retrospective study that utilized aggregated monthly TB cases among children data reported by health facilities located in Homa Bay and Turkana Counties to the National Tuberculosis, Leprosy and Lung Disease Program (NTLLDP) in the Treatment Information from Basic Unit (TIBU) electronic system between January 2012 to December 2021. The data comprised 120 data points. The study utilized data reported by health facilities in Homa bay and Turkana Counties which are among the top 10 TB endemic Counties in Kenya [50].

### Data collection and analysis

Tuberculosis case data were abstracted and aggregated for each month between January 2012 to December 2021 for health facilities located in Homa bay and Turkana Counties in Kenya. In 2012, the Kenya Ministry of Health (MoH) through the Division of Leprosy, Tuberculosis and Lung Disease transitioned the reporting of TB cases from paper-based to the TIBU system [23]. The TIBU system is a national case-based surveillance system used in the storage of individual cases of TB that are reported to the national TB program monthly with nationwide coverage [24]. This study did not collect or utilize patient-level data.

The 120 TB cases data points in the data used in this study were split using the 80:20 ratio with 80% (2012 to 2019) of the data assigned to the training set and 20% (2020 to 2021) assigned to the testing set. Splitting the data into training and testing data using the 80:20 ratio, in this case, was based on the available chronological data points as this has been proved to yield test error rate estimates with reasonably low bias and variance [25]. Furthermore, when the training data is large enough, the model learns well and this gives predicted values that are much closer to the actual values [26]. The models were built on the training set and their performance was evaluated on the testing set; this method is the holdout-validation method for model performance evaluation.

Data analysis was performed using R statistical software [27] together with applicable packages for analyzing time-series data. The results were summarized using tables and figures.

### The Time Series concept

A time series is a sequential set of data points measured over time and is typically composed of the trend, cyclical, seasonal, and irregular (random) components.

An autoregressive (AR) model is a type of random process used to describe certain time-varying processes within a time series [28]. The basic idea of AR models is that the present value of a series Y_t_ can be linearly explained by a function of *p* past values, that is,Y_t−1_, Y_t−2_ +, …, + Y_t−p_.

Assuming, E(Y_t_) = 0, an AR process of order *p* can be written as;

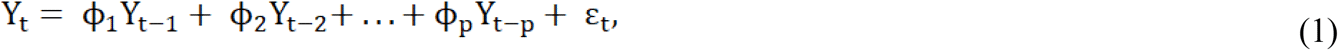

Where ε_t_ is white noise (WN), and is uncorrelated with Y_s_ for all s < t

However, if the mean is E(Y_t_) = µ ≠ 0, then Y_t_ is replaced by Y_t_ − µ to obtain;

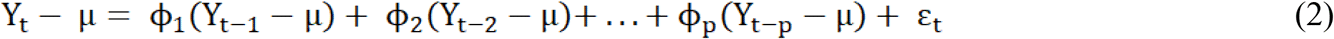

Equation 2 can also be written as;

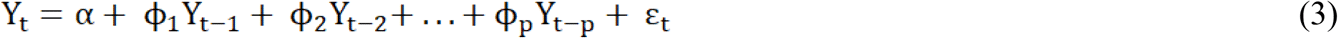

Where;

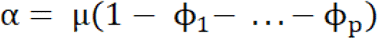

Furthermore, equation (1) can be written in the form;

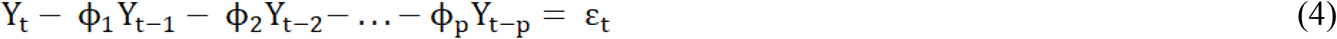

However, by applying the backshift operator BY_t_ = Y_t−1_, we get;

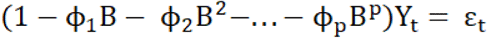

Or using notation, we can write;

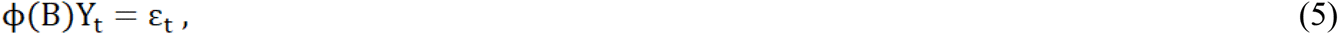

Where ϕ(B) denotes the autoregressive (AR) operator;

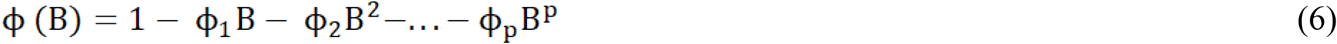

A moving average (MA) model uses the dependency between an observed value and the residual error from a moving average model applied to the lagged observations. This implies that the output variable is linearly dependent on the current and past values of a stochastic term [28]. Consequently, Y_t_ is a moving average process of order *q* if

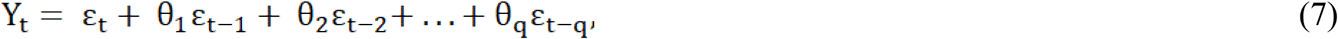

Where ε_t_ is WN and θ_1_, …, θ_q_ are constants

On the other hand, a MA(*q*) process can also be written in the form;

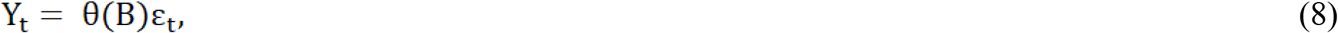

Where the moving average operator;

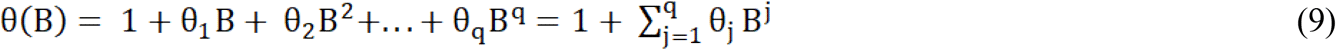

defines a linear combination of values in the shift operator B^k^ε_t_ = ε_t−k_

### Autoregressive Integrated Moving Average models (ARIMA) models

An ARMA (*p, q*) model is a class of stochastic processes whose auto-covariance functions depend on a finite number of unknown parameters. Generally, an ARMA process of orders *p* and *q* can be represented mathematically [29] as;

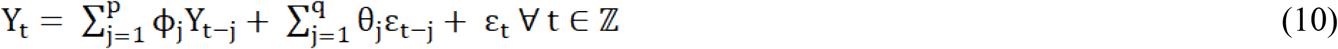

In the lag operator notation, the ARMA (*p, q*) process is given by

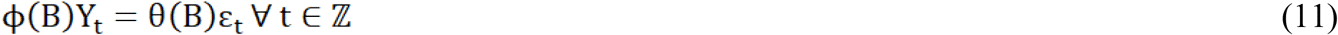

Box and Jenkins introduced the ARIMA model in 1960 [30]. The ARIMA model requires only historical time series data on the variable under forecasting. Most importantly, ARIMA models are represented as ARIMA (*p, d, q*) where *p* is the number of AR terms, *d* is the number of non-seasonal differences, and *q* is the number of lagged forecast errors [31]. The ARIMA model assumes that the residuals are independent and normally distributed with homogeneity of variance and zero mean value.

### Seasonal Autoregressive Integrated Moving Average models (SARIMA) models

The SARIMA model is made up of non-seasonal and seasonal components in a multiplicative model. A SARIMA model can be written as ARIMA (*p,d,q*) (*P, D, Q*)*S* where *p* is the non-seasonal AR order, *d* is the non-seasonal differencing, *q* is the non-seasonal MA order, *P* is the seasonal AR order, *D* is the seasonal differencing, *Q* is the seasonal MA order and *S* is the period of repeating seasonal pattern. Generally, S=12 for monthly data.

Let the backshift operator be presented as *B*Y_t_ = Y_t-1_

Without differencing, a SARIMA model can be written formally as;

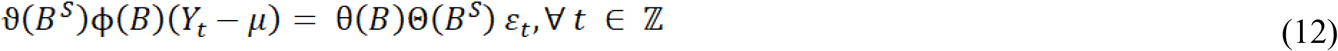

Where on the left of equation 12, the seasonal and non-seasonal AR processes multiply each other, and on the right of equation 12, the seasonal and non-seasonal MA processes multiply each other. Also, in this study, S=12 since monthly TB case data are used.

### Artificial Neural Networks (ANNs) models

Artificial Neural Networks have been suggested as alternative and better modeling approaches to time series forecasting [32]. The main goal of ANNs is to construct a model that mimics the human brain intelligence into a machine [33] and is biologically motivated [34]. The most common ANNs are multi-layer perceptrons (MLPs) that utilize a single hidden layer feed-forward network (FNN) [35] made up of the input layer, the hidden layer, and the output layer connected by acyclic links [36]. A neuron is a data processing unit while the nodes in the various layers of ANNs are the processing elements. In this study, the inputs were the lagged TB case observations and the outputs were the predicted or fitted TB cases from the model. According to Zhang [37], the ANN can be written as:

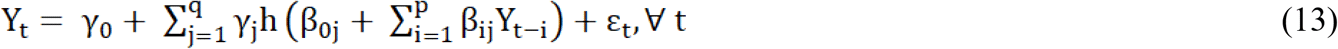

Where Y_t-i_ *(i=1, 2, …, p)* are the *p* inputs, Y_t_ is the output, *q* are the hidden nodes, γ_j_*(j=0, 1, 2, …, q)* and β_ij_ *(i=0, 1, 2, …, p; j=0, 1, 2, …, q)* are the connection weights and ε_t_ is the random shock; γ_0_ and β_0j_ are the bias terms. There is no systematic rule in deciding the choice of *q* while *p*, which is the number of neurons is equal to the number of features in the data. The logistic function *h(*.) is applied as the nonlinear activation function, where:

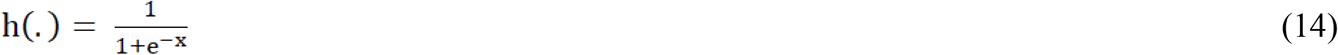

As a matter of fact, the model in equation 13 performs a nonlinear functional mapping from past observations of a time series to the future value. That is:

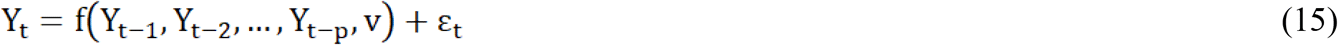

Where *v* is a vector of all parameters and f(.) is a function determined by the structure of the network and the connection weights.

### Hybrid (ARIMA-ANN) models

Generally, a time series can be observed as having linear and nonlinear components as shown in equation 16.

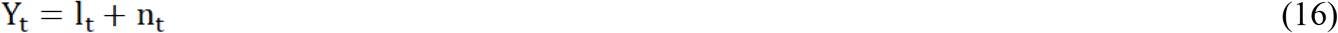

Where l_t_ and n_t_ are the linear (from the ARIMA model) and nonlinear (ANN fitted ARIMA model residuals) components respectively. Residuals from the ARIMA model are fitted with the ANN model.

### Proposed Methodology

The proposed methodology for this study was based on the combination of the Box-Jenkins methodology for ARIMA modeling, and the ANN and hybrid ARIMA models as shown in Fig 1.

**Fig 1:**
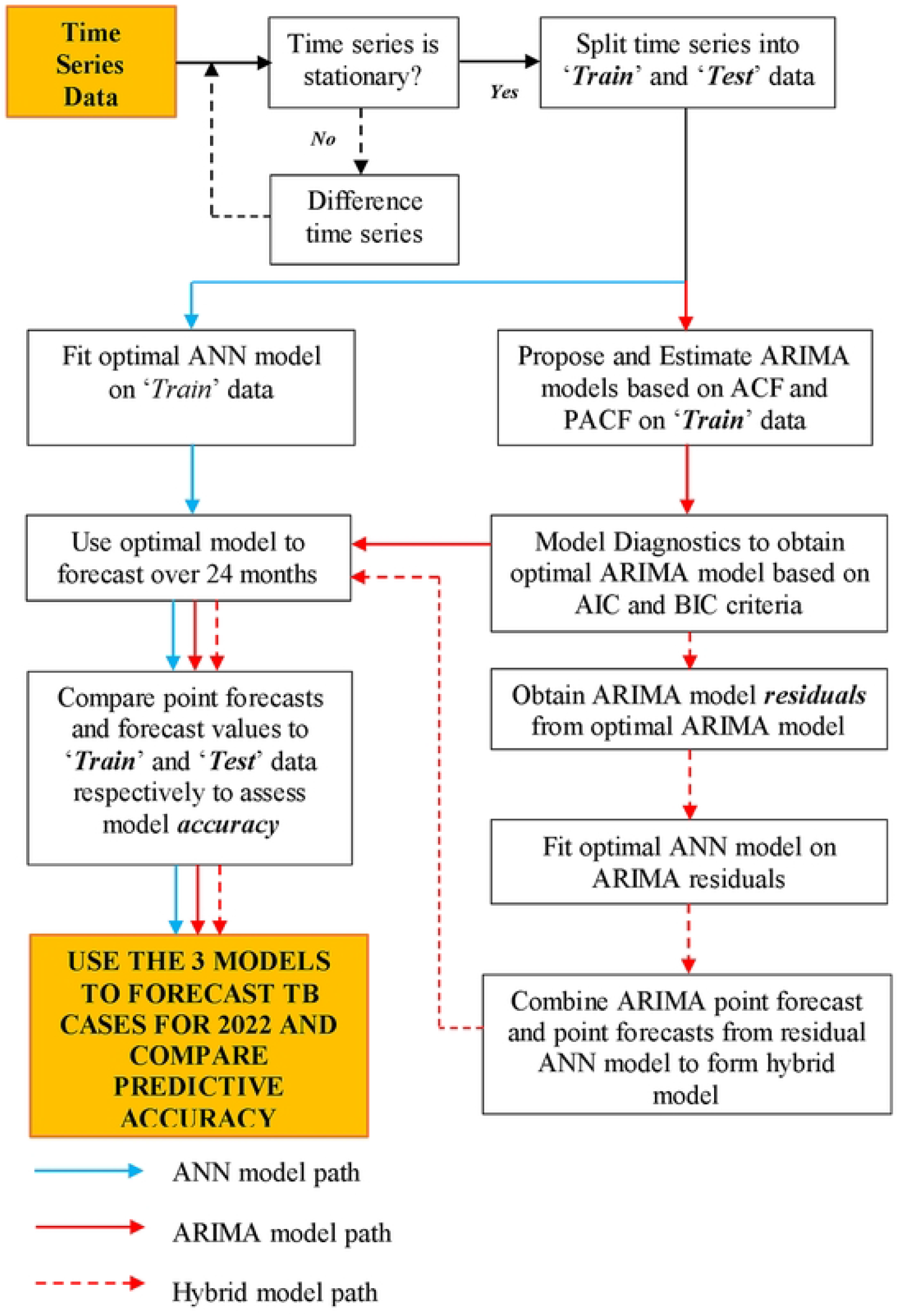
The Proposed methodology.

### Model identification and specification

Optimal values of *p, d*, and *q* for the ARIMA model were determined by examining the autocorrelation functions and the best model was determined by testing models with different parameters of *p, d*, and *q*. The models were estimated using the maximum likelihood estimation (MLE) method and the Akaike Information Criterion (AIC) and Bayesian Information Criterion (BIC) [38] penalty function statistics were used to determine the best model that minimizes AIC or BIC.

One assumption of the ARIMA model is that the residuals should be white noise. As such, the Ljung-Box Q test [39] was used to test the hypothesis of independence, constant variance and zero mean of the model residuals.

### Accuracy measures

Various accuracy measures have been proposed [40] to determine predictive and forecast performance. This study used the Root Mean Squared Error (RMSE), Mean Absolute Error (MAE), and the Mean Absolute Percent Error (MAPE) to measure the predictive and forecast accuracy of the three models. The lower the values of these accuracy measures the better the model. Furthermore, MAPE values of 10% or below, 10-20%, and 20-50% should be considered as high accuracy, good accuracy, and reasonable accuracy [41].

The study also compared the predictive accuracy of the forecasts from the three models using the Diebold-Mariano (DM) test [42]. The test was used to test the null hypothesis that two models have similar predictive accuracy.

### Ethical approval and considerations

Authorization for use of the data from the TIBU system was obtained from the National Commission for Science, Technology, and Innovation (NACOSTI) through a research permit and letters of approval from the research unit of the department of health services Homabay County and the department of medical services Turkana County.

## Results

### Exploratory data analysis

There was a total of 120 data points in this dataset. The trend of the TB cases among children below 15 years in Homa bay and Turkana counties in the data is shown in Fig 2 showing a marginal increase in the TB cases reported between 2018 and 2021.

**Fig 2:**
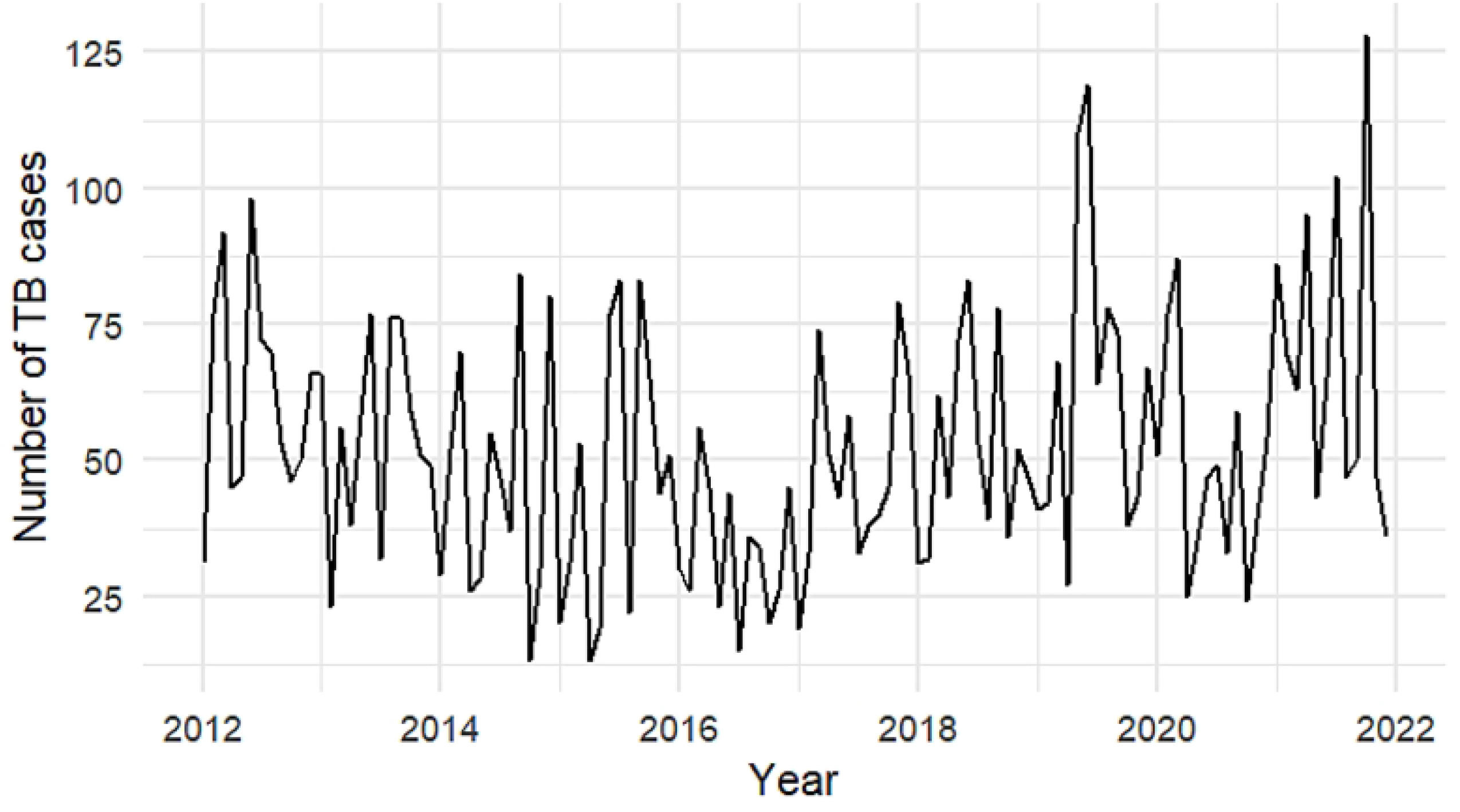
Monthly TB cases among children below 15 years from Homabay and Turkana Counties between 2012 and 2021.

The monthly cycle box plot of TB cases is shown in Fig 3 where there is a potential presence of seasonality within the reported TB cases. However, whether or not to account for seasonality in the model depends on whether this would improve model accuracy. This implies that there is need to account for seasonality within the ARIMA model. Furthermore, outliers were detected in some months.

**Fig 3:**
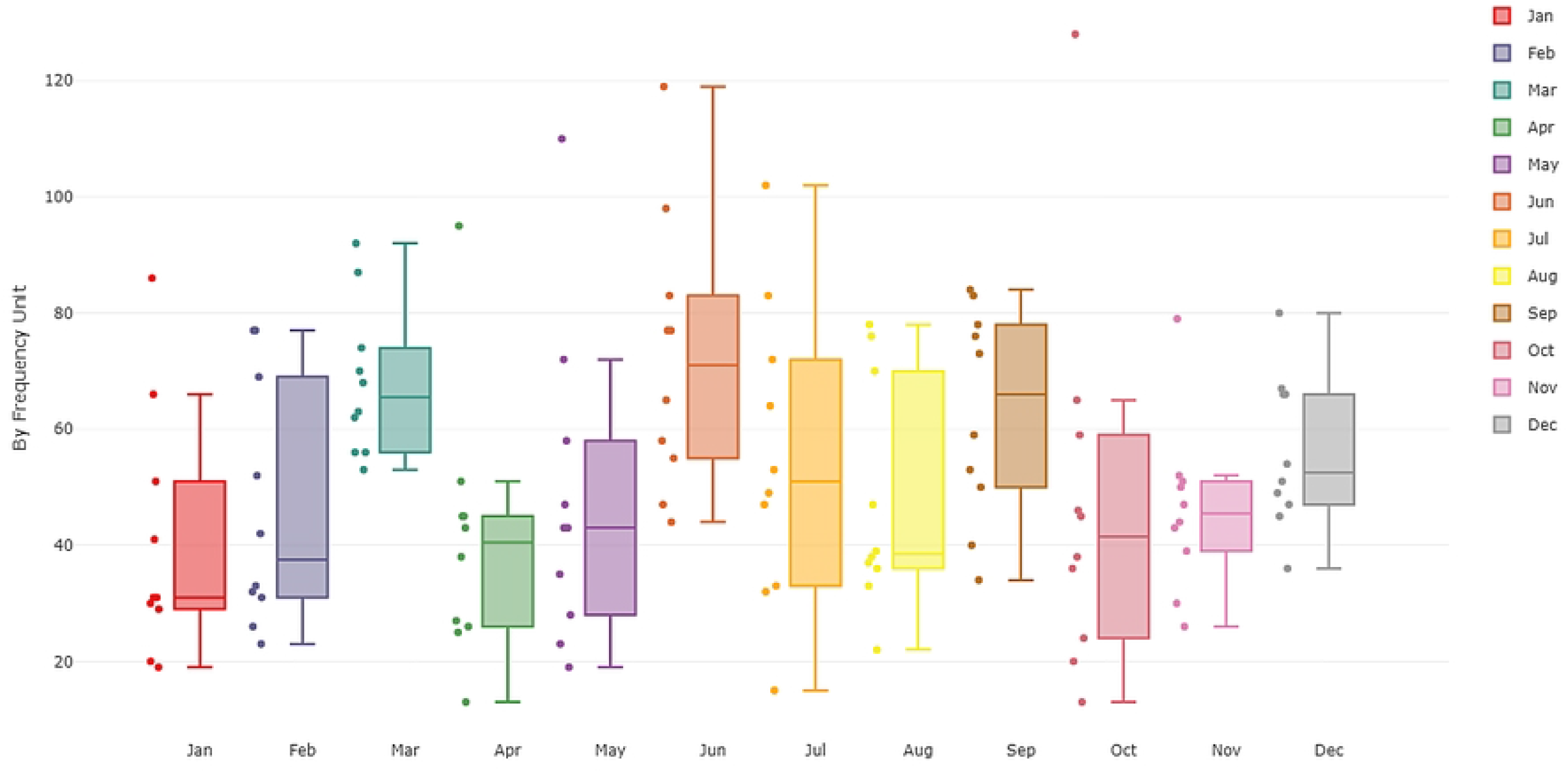
Monthly cycle plot of TB cases.

### Comparison of model performance in predicting TB cases among children below 15 years in Homabay and Turkana Counties, Kenya

#### ARIMA Model estimation and accuracy

The Akaike Information Criterion (AIC) and the Bayesian Information Criterion (BIC) were used to pick the best parsimonious model based on the least AIC or BIC estimated values. The best model was ARIMA (0,0,1,1,0,1,12); where *p*=0, *d*=0 and *q*=1 respectively and *P*=1, *D*=0 and *Q*=1 respectively. The Ljung-Box Q test for the best model showed a p-value of 0.971 implying that the ARIMA (0,0,1,1,0,1,12) model residuals were independently distributed.

The best model was made up of non-differenced seasonal AR (1), non-seasonal MA (1) model and seasonal MA (1) polynomials and using equation 12, the ARIMA (0,0,1,1,0,1,12) model equation can be written as:

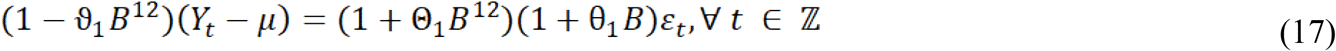

Where is ε_t_ WN, and uncorrelated with Ys for each s < t

From the model output, the estimated coefficients were (see Table 1); ma1 = θ_1_ = 0.291, sar1 =ϑ_1_ = 0.997, sma1 = Θ_1_ = -0.953 and *µ* = Intercept = 50.902

**Table 1:** Estimated model coefficients.

|  | Coefficients | Std. Error | Z-value | Pr(>\|z\|) |
| --- | --- | --- | --- | --- |
| ma1 | 0.291 | 0.108 | 2.701 | 0.007* |
| sar1 | 0.997 | 0.015 | 68.167 | <0.001** |
| sma1 | -0.953 | 0.127 | -7.512 | <0.001** |
| Intercept | 50.902 | 5.064 | 10.052 | <0.001** |

Significance codes: 0 '\*\*\*' 0.001 '\*\*' 0.01 '\*' 0.05 '.' 0.1 ' ' 1

Plugging these estimated coefficients into equation 17, yields the model equation:

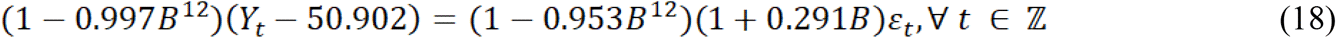

### ARIMA Model residual diagnostics

The best ARIMA model was assessed for fit using the standard model residual analysis. In model diagnostic checking, 4 plots were used to test the underlying assumptions as shown in Fig 4. The Q-Q plot was relatively normal except for a few outliers at the tails, with model residuals being normally distributed. Inspection of the Autocorrelation Function (ACF) and Partial Autocorrelation Function (PACF) plots to test residual randomness to identify patterns or extreme values showed significant auto-correlations at lag 3.

**Fig 4:**
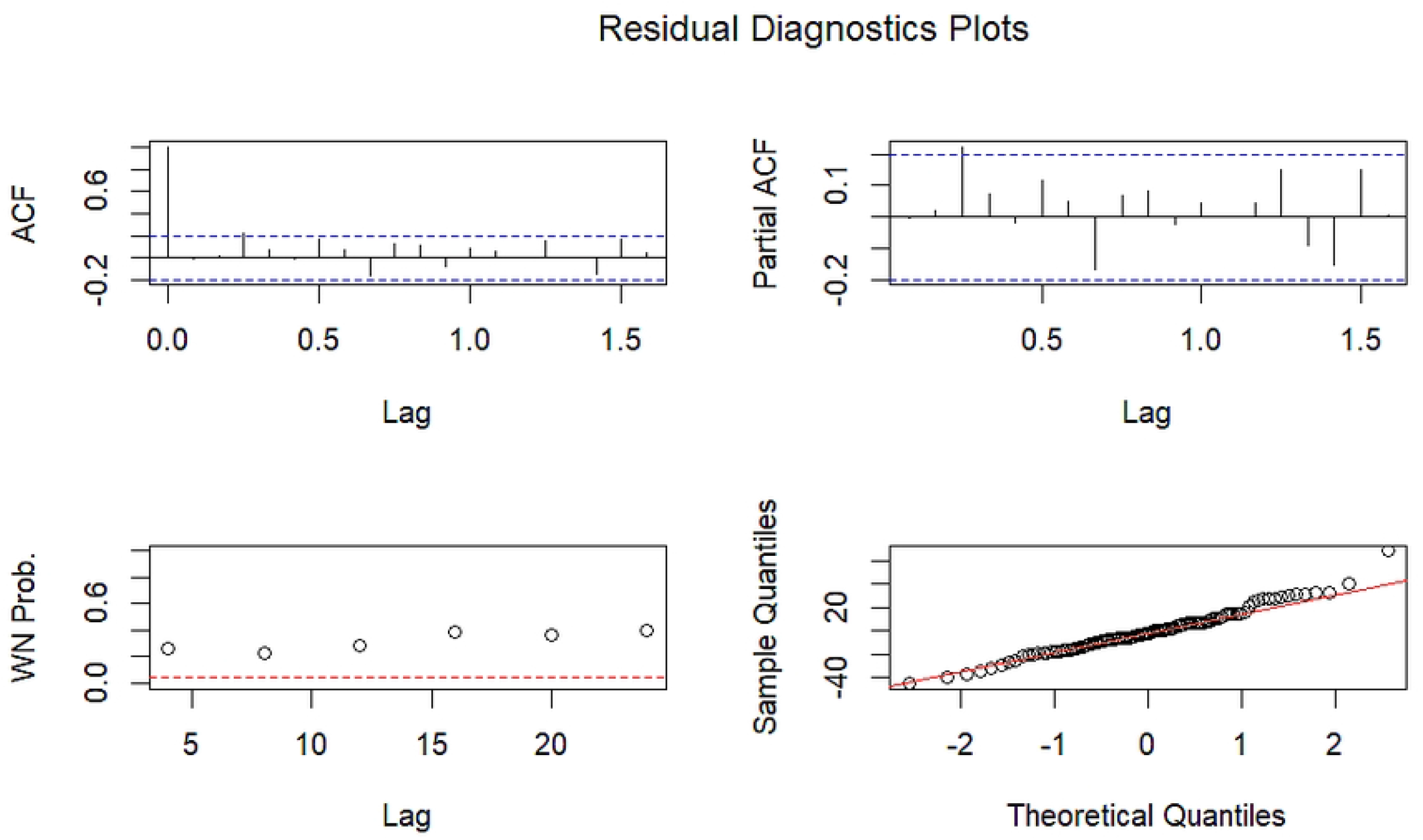
ARIMA Model residual diagnostics.

### ARIMA model performance

The performance of the ARIMA (0,0,1,1,0,1,12) model was carried out by comparing predicted and forecasted TB cases with the actual TB cases reported and presented in Fig 5 and Fig 6 respectively.

**Fig 5:**
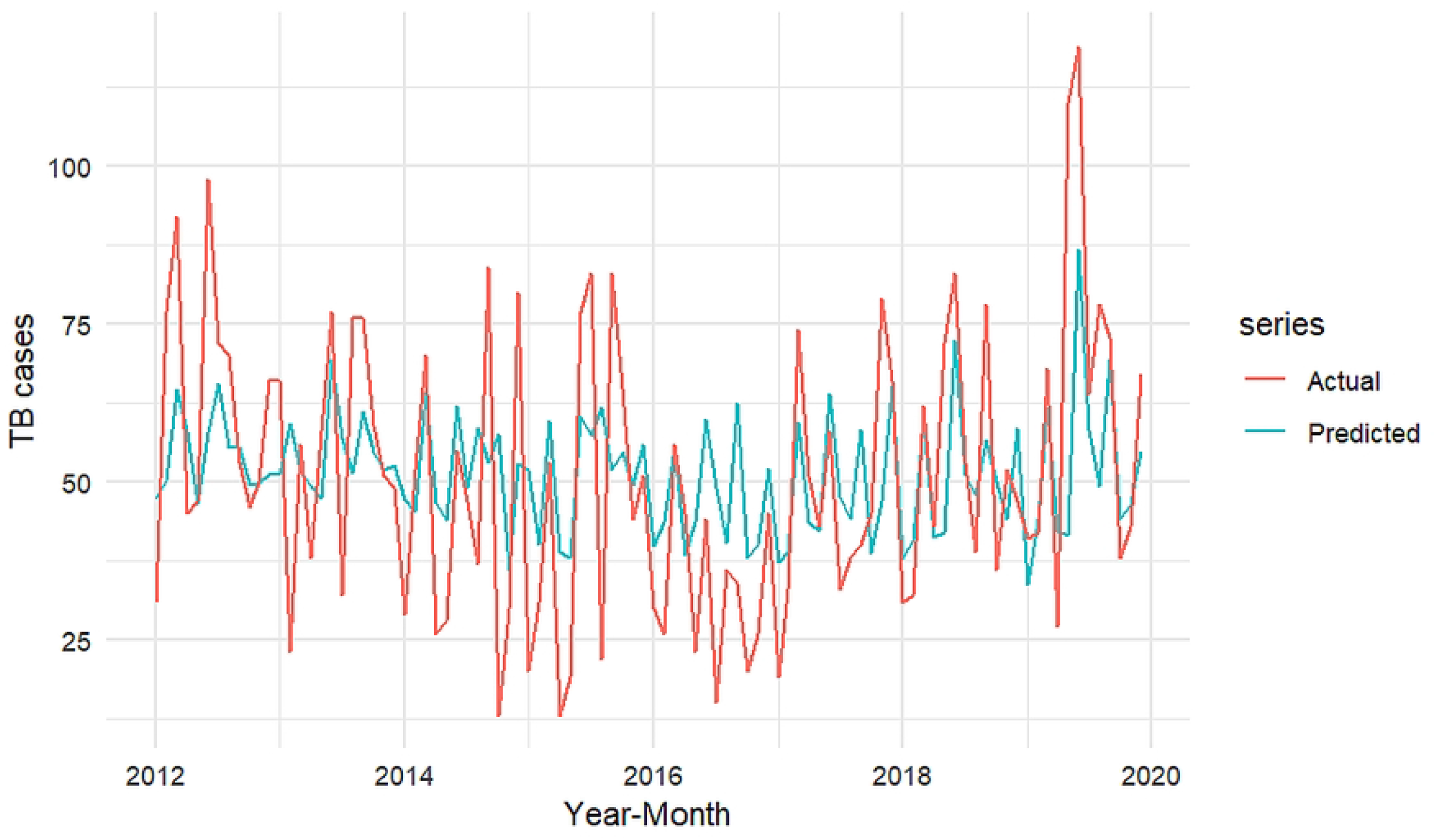
Comparison of ARIMA predicted versus actual (training data) TB cases.

**Fig 6:**
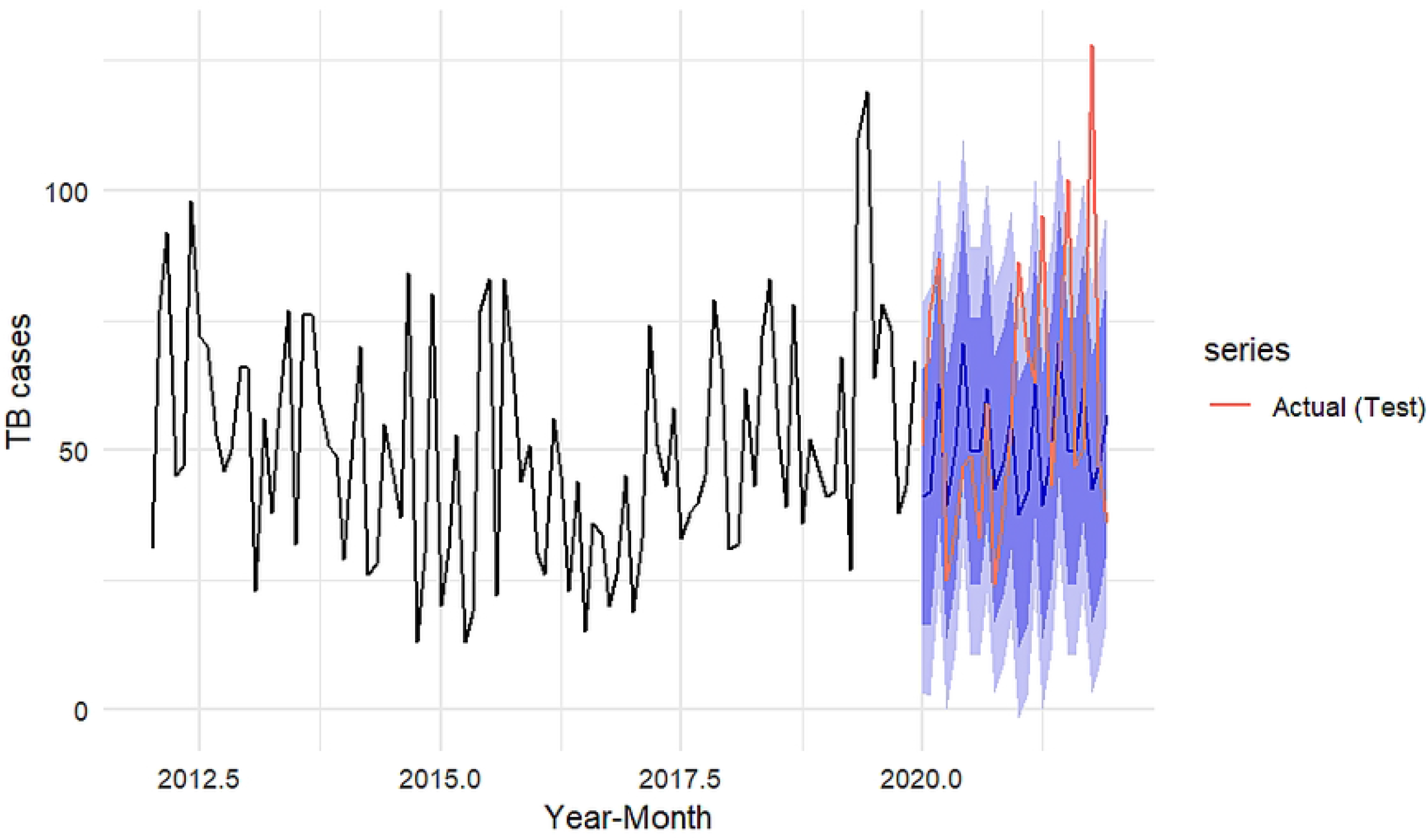
Comparison of ARIMA 24-month forecast versus actual (test data) TB cases.

### ARIMA (0,0,1,1,1,0,1,12) model accuracy

Table 2 compares the accuracy of parameters/measures of the ARIMA (0,0,1,1,0,1,12) model on the training (2012 to 2019) and testing (2020 to 2021) data. The accuracy measures compared were RMSE, MAE, and MAPE correspondingly.

**Table 2:** Model (ARIMA (0,0,1,1,0,1,12)) accuracy comparison on training and testing data.

| Data | RMSE | MAE | MAPE |
| --- | --- | --- | --- |
| Training | 18.74 | 14.39 | 39.00 |
| Testing | 29.17 | 20.47 | 33.15 |

The accuracy measure comparison in table 2 demonstrates that the model performs slightly worse on the testing data an RMSE value of 29.17 compared to 18.74 on the training data.

Large RMSE values generally imply that the fitted model fails to account for important information within the underlying data hence the need to account for such information which might be captured within the non-linearities as shown in the additive hybrid model methodology which provides fitted values that are as closer to the actual/observed data.

### Artificial Neural Network (ANN) model estimation and accuracy

The training dataset was fit using an ANN model using the Neural Network Auto-Regressive (NNAR) function to produce an NNAR (*p,P,k*) [m] model. The optimal lag parameter, *p*, and the number of nodes in the hidden layer, *k*, were automatically selected while *P*=1 by default. In addition, a decay parameter of 0.001 and a maximum iteration of 200 were pre-set for the model to help restrict the weights from becoming too large and ensure that the model can test different models until the optimal model that has the minimal RMSE produced respectively.

The optimal NNAR model produced was NNAR (1,1,2) [12] with an average of 20 networks each of which was a 2-2-1 network with 9 weights. The plot of the point forecast values from the NNAR (1,1,2) [12] model on the training set against the actual training data were presented in Fig 7 while Fig 8 presents the 24-month forecast based on the NNAR (1,1,2) [12] model. Table 3 shows the accuracy measures from the model. The model seems to perform badly on the testing dataset although the MAPE values below 50% are reasonable.

**Table 3:** Model (NNAR (1,1,2) [12]) accuracy comparison between training and testing set.

| **Data** | **RMSE** | **MAE** | **MAPE** |
| --- | --- | --- | --- |
| Training | 18.56 | 14.58 | 29.89 |
| Testing | 28.65 | 21.95 | 38.86 |

**Fig 7:**
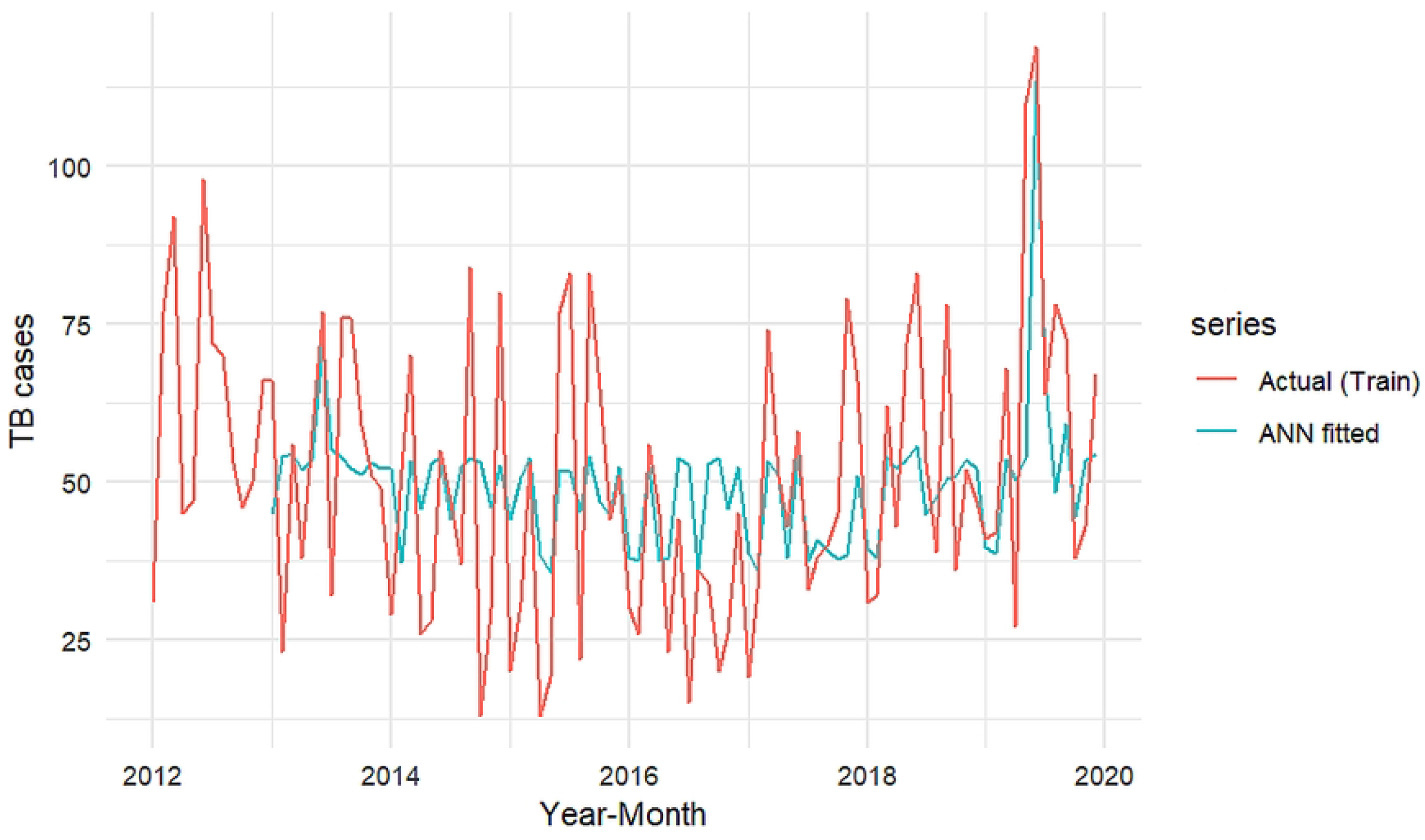
Comparison of NNAR (1,1,2) [12] predicted TB cases versus actual TB cases (training data)

**Fig 8:**
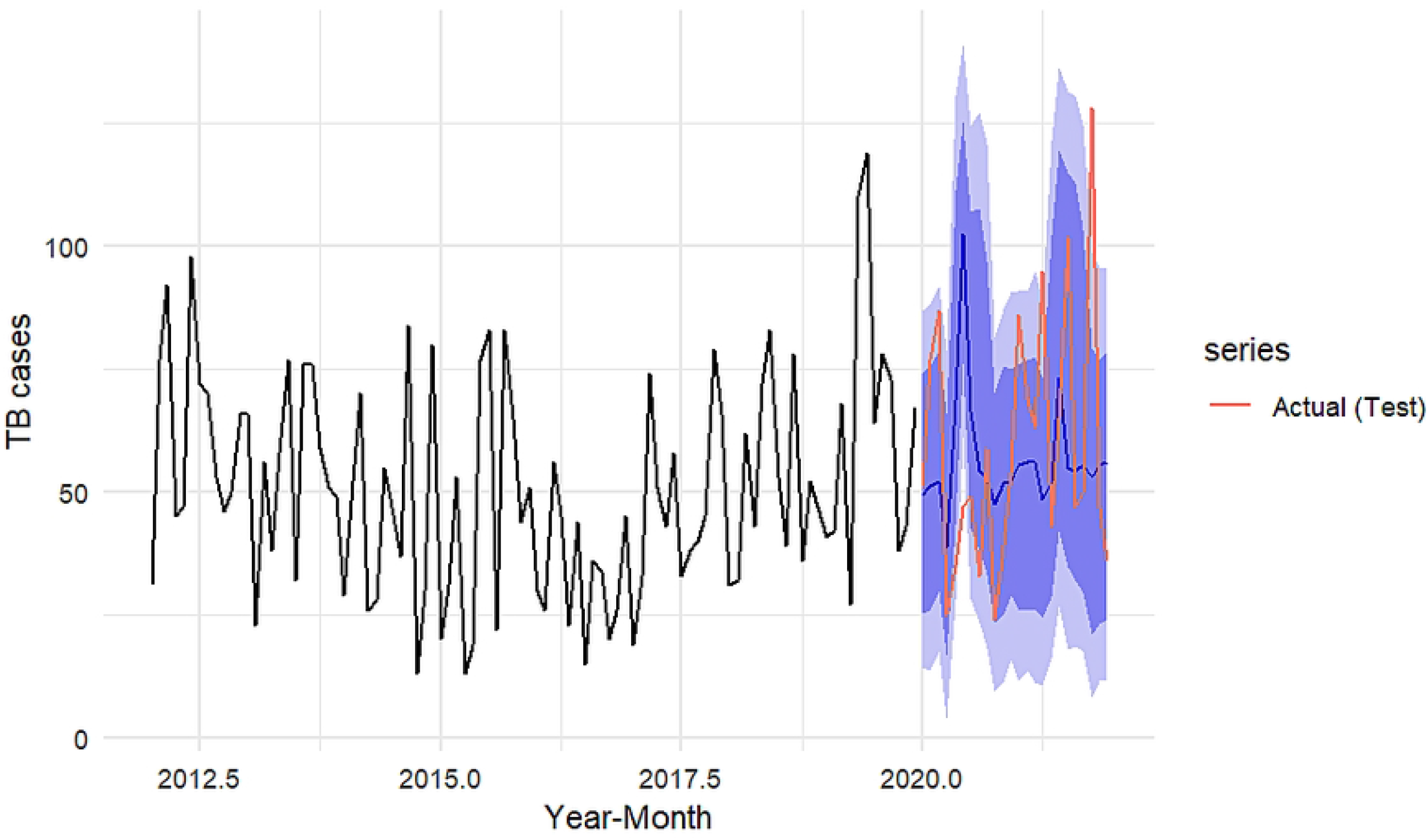
Comparison of NNAR (1,1,2) [12] 24-month forecast TB cases versus actual TB cases (test data)

### Hybrid (ARIMA-ANN) model estimation and accuracy

Residuals from the optimal ARIMA (0,0,1,1,0,1,12) model were fit using an ANN model and the accuracy measures and comparison of the forecast and prediction were presented in Table 4, Fig 9 and Fig 10 respectively.

**Table 4:**
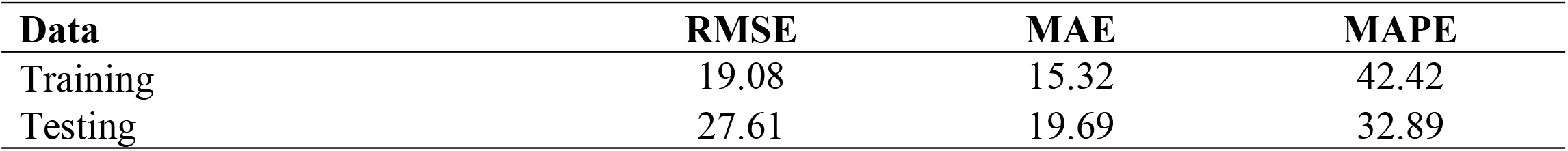
Hybrid ARIMA-ANN model accuracy.

| **Data** | **RMSE** | **MAE** | **MAPE** |
| --- | --- | --- | --- |
| Training | 19.08 | 15.32 | 42.42 |
| Testing | 27.61 | 19.69 | 32.89 |

**Fig 9:**
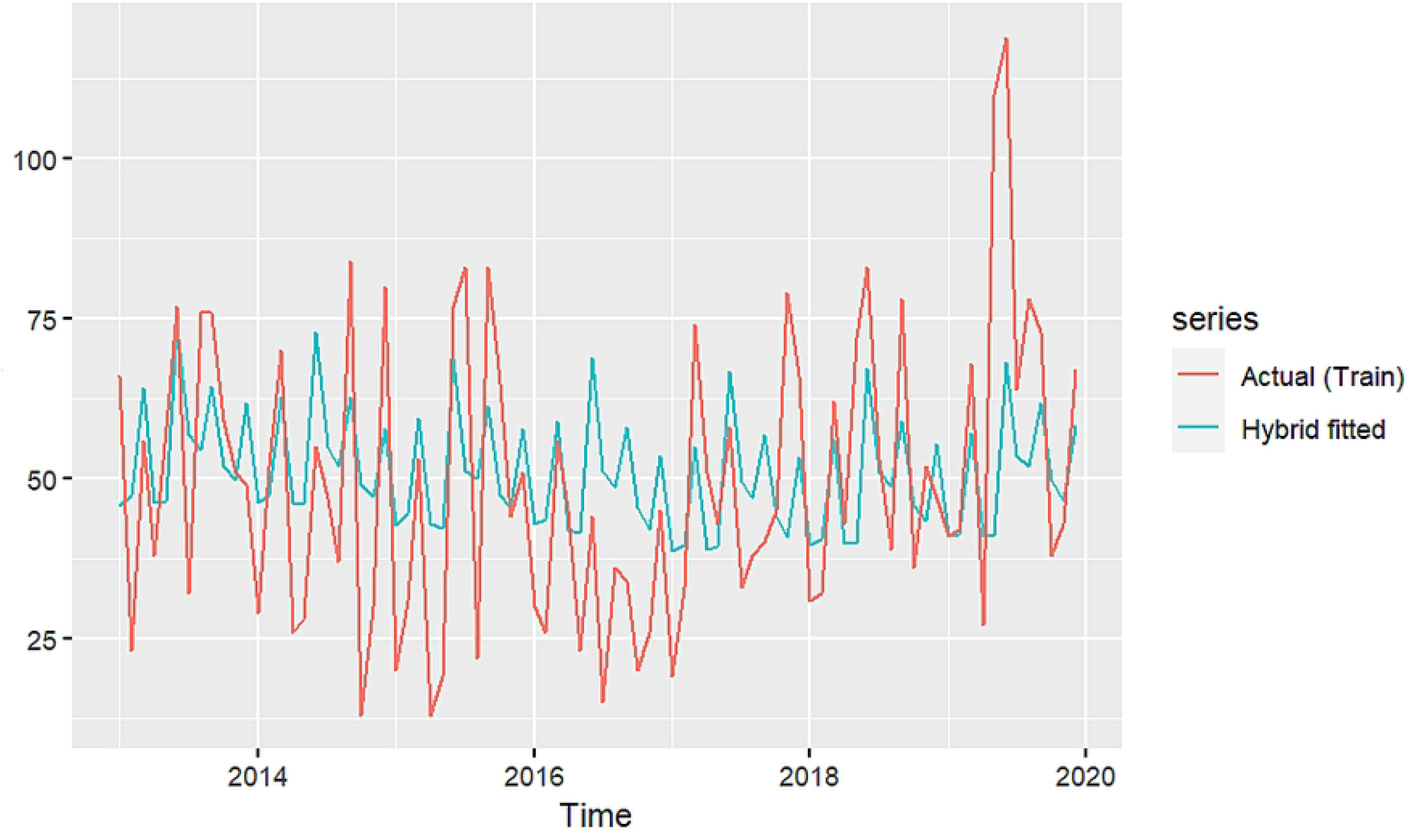
Comparison of ARIMA-ANN predicted TB cases versus actual TB cases (training data)

**Fig 10:**
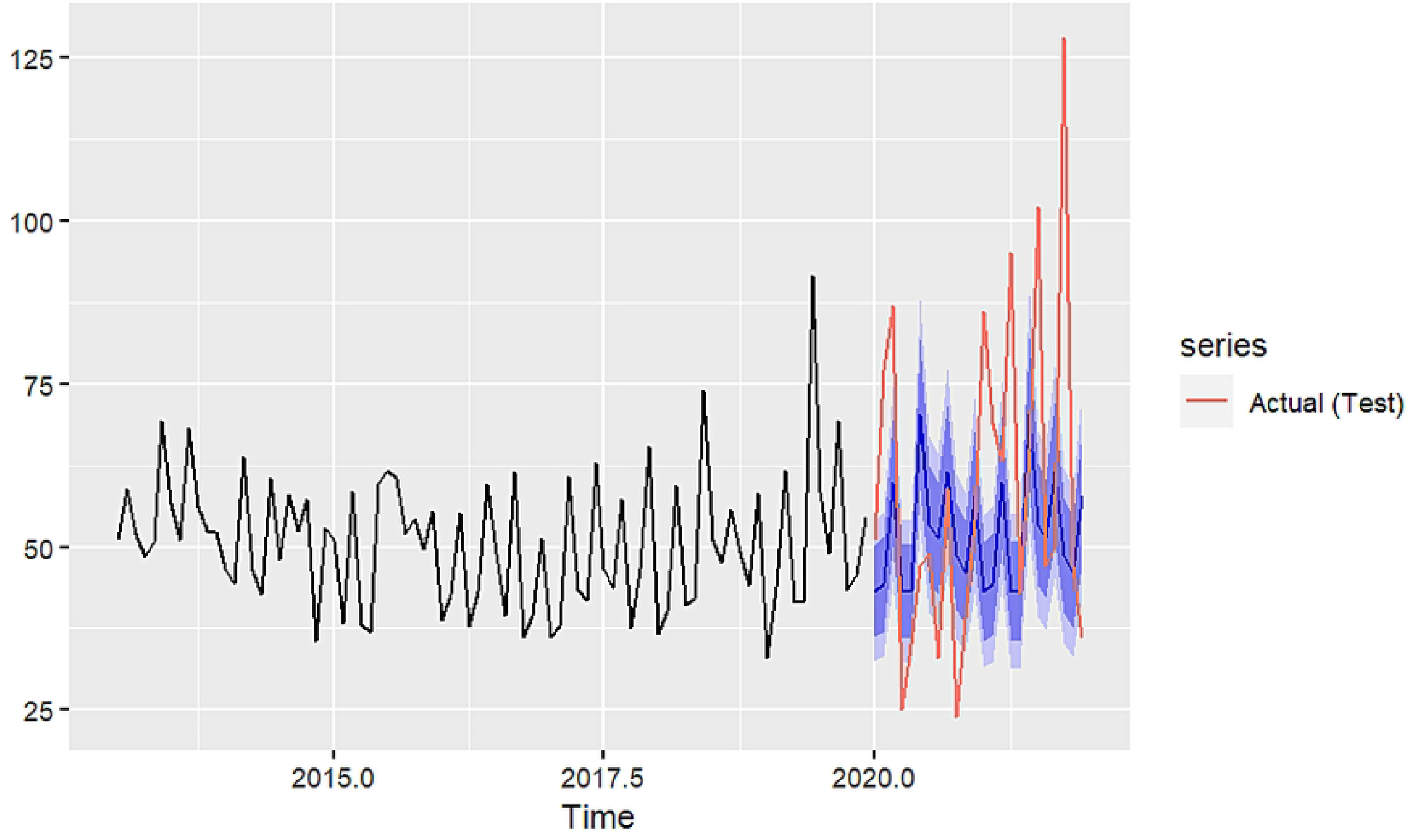
Comparison of ARIMA-ANN 24-month forecasted TB cases versus actual TB cases (testing data)

### Comparison of predictive accuracy of the models

The predictive accuracy of the models was compared using the Diebold-Mariano (DM) test with the null hypothesis that the predictive accuracy of the two models compared are the same. The results in table 5 show that the NNAR (1,1,2) [12] and ARIMA-ANN models present significantly different predictive accuracies, similar to ARIMA (0,0,1,1,0,1,12) versus ARIMA-ANN models. In general, the ARIMA-ANN model offers better predictive accuracy compared to the NNAR (1,1,2) [12] and ARIMA (0,0,1,1,0,1,12) models.

**Table 5:** Comparison of predictive accuracy.

| Model | DM statistic | Loss Function Power | P-value |
| --- | --- | --- | --- |
| ARIMA (0,0,1,1,0,1,12) Vs NNAR (1,1,2) [12] | 0.732 | 2 | 0.466 |
| NNAR (1,1,2) [12] Vs ARIMA-ANN | 6.260 | 2 | <0.001 |
| ARIMA (0,0,1,1,0,1,12) Vs ARIMA-ANN | 8.732 | 2 | <0.001 |

### Comparison of model performance in forecasting temporal trends of TB incidences

The resulting ARIMA (0,0,1,1,0,1,12), NNAR (1,1,2) [12] and ARIMA-ANN models were used to forecast TB cases for the next 12 months (2022). The forecast results were presented in table 6. The 12-month mean forecasted TB cases was 55, 59 and 52 cases per month based on the ARIMA (0,0,1,1,0,1,12), ANN (1,1,2) [12] and ARIMA-ANN respectively for 2022 giving a total of 657, 706 and 629 TB cases forecasted for the year 2022 from the ARIMA (0,0,1,1,0,1,12), ANN (1,1,2) [12] and ARIMA-ANN models respectively.

**Table 6:** TB cases 12 month point forecast comparison.

| Month | ARIMA (0,0,1,1,0,1,12) | NNAR (1,1,2) [12] | ARIMA-ANN |
| --- | --- | --- | --- |
| Jan-22 | 57 | 53 | 44 |
| Feb-22 | 61 | 53 | 45 |
| Mar-22 | 62 | 52 | 60 |
| Apr-22 | 56 | 58 | 43 |
| May-22 | 46 | 52 | 44 |
| Jun-22 | 54 | 52 | 71 |
| Jul-22 | 63 | 72 | 54 |
| Aug-22 | 46 | 52 | 52 |
| Sep-22 | 53 | 56 | 62 |
| Oct-22 | 63 | 103 | 49 |
| Nov-22 | 48 | 57 | 46 |
| Dec-22 | 48 | 46 | 59 |
| **Mean** | **55** | **59** | **52** |
| **Total** | **657** | **706** | **629** |

## Discussion

Although all three models were able to predict TB cases among children below 15 years, the hybrid ARIMA-ANN model was able to offer better predictive performance compared to single ARIMA and ANN models. These findings compare with those from other studies which applied either hybridized ARIMA or SARIMA in the modeling of TB incidences and other infectious diseases [12, 16, 43, 44] with the overall conclusion that hybrid models have better predictive performance. The majority of infectious disease data are neither purely linear nor non-linear and mostly present with both linear and nonlinear properties. As such, single models are not enough in modeling such kinds of data. Hybrid models are found to be most appropriate for the accurate estimation of such data [45]. The use of hybridized ARIMA models has been proposed in recent years and used extensively with improvements proposed over time.

The estimated TB prevalence in Kenya was 259 TB cases per 100,000 population in 2020 [4]. This translates to approximately 134,680 TB cases and with children accounting for about 20% (26,936) of these cases [46], the prevalence among children below 15 years was approximately 121 TB cases per 100,000 population of children.

From the forecasted mean of 52 to 59 TB cases per month in 2022 for Homabay and Turkana Counties, this study estimates that for the year 2022, the number of TB cases reported would be approximately 629 to 706. However, given that these are estimated reported cases, they most likely represent only about 35% of TB cases since up to 65% of pediatric TB cases are potentially missed each year [2]. Taking this into account, the estimated TB cases for 2022 will be approximately 1797 to 2017 for Homa Bay and Turkana Counties among children. The estimated population of children below 15 years in Homabay and Turkana Counties for 2022 is approximately 1,020,795 [47]. As such, the estimated TB prevalence among children in Homa Bay and Turkana Counties in 2022 would be approximately 176 to 198 TB incidences per 100,000 population. These TB prevalence estimates for 2022 are way higher than the estimated national average of 121 per 100,000 population of children below 15 years in 2020.

The findings of this study show that the estimated TB prevalence among children below 15 years is much higher compared to the estimated national average for 2020. These findings are in line with the WHO newsletter that showed that the number of people developing TB and dying from the disease could be much higher in 2021 and 2022 mainly because of the COVID-19 pandemic [48]. These findings also confirm those by Oliwa *et al*. who indicated that notification data may underestimate the TB burden among children [49] while Mbithi *et al*. reported a decrease in TB diagnosis in Kenya by an average of 28% in the year 2020 [50].

## Conclusion

The hybrid ARIMA model offers better predictive accuracy and forecast performance compared to single ARIMA and ANN models in modeling TB cases among children below 15 years in Homabay and Turkana Counties.

The findings in this study allude to the under-reporting of TB cases among children below 15 years. As such, there is need to re-look at the TB surveillance framework data more closely to understand existing gaps. There is an urgency to re-align vital resources towards the National TB program to have the TB fight back on track.

## Limitations

This study utilized data collected and reported in the TIBU system, as such, the study did not have control over the quality and accuracy of the data.

This study utilized data between 2012 to 2021 which comprised 120 data points. Deep learning algorithms such as ANNs usually demand a large amount of data to allow the algorithm to effectively learn. In this study, there were 96 data points for model development and while this represented 80% of the records, it might not have been sufficient enough to allow proper learning of the algorithm. Furthermore, there were only 24 records used for model validation and this might not have been large enough to allow for better learning by the algorithm.

This study combined data and analysis for Turkana and Homabay County. However, these two counties might present different scenarios when it comes to pediatric TB.

Finally, since the study focused on modeling TB cases among children below 15 years in Homa bay and Turkana counties, the findings might not be generalized to other counties of Kenya.

## Data Availability

Yes - all data are fully available without restriction. For the purposes of this study, all of the data necessary to replicate the analysis are contained in the tables and figures presented in the manuscript. If this is deemed not to be the case for any researchers wishing to conduct secondary analyses or if supplementary materials are needed, requests may be submitted through me as the corresponding author. I will ensure reasonable requests are granted.

## Acknowledgments

We would like to thank the departments of health in Homa bay and Turkana County for granting permission to access and utilize TB data in the TIBU system, and we hope that this work aids in the realization of the goals of the TB program in these two counties and beyond.

## Notes

### Competing Interest Statement

The authors have declared no competing interest.

### Funding Statement

The study was personally funded as part of the requirement to graduate for a Master of Science program at the University of Eldoret. The funder had a role in study design, data collection and analysis, decision to publish, and preparation of the manuscript.

### Author Declarations

Research permit is applied and received from the National Commission for Science, Technology and Innovation in Kenya Approval is also applied and received from the departments of health from Homa bay and Turkana Counties, Kenya

